# Preventing disproportionate mortality in ICU overload situations: Empirical evidence from the first COVID-19 wave in Europe

**DOI:** 10.1101/2021.05.03.21255735

**Authors:** Paul Buijs, Rodolfo Catena, Matthias Holweg, Taco van der Vaart

## Abstract

Avoiding overloading the healthcare system remains a central issue during the COVID-19 pandemic. The logic of preventing such overload situations is intuitive since the level and quality of critical care is a function of the available capacity to provide it. Where this capacity is no longer available due to a surge in admissions, patient outcomes will invariably deteriorate in the long run – ultimately leading to disproportionate mortality. In this paper, we study the three worst affected regions in Italy, the Netherlands, and Germany during the first COVID-19 wave in the spring of 2020. We report on quantitative analyses that show how mortality rises non-linearly as the proportion of COVID-19 patients in the ICU increases. We identify changes to the patient-staff ratio, increasing exhaustion and infection levels amongst staff, as well as equipment shortages, as likely causes driving this rise in mortality. We explore these findings further with interviews of key stakeholders in the respective healthcare systems. Our results demonstrate that the common approach of managing COVID-19 surges by stretching ICU capacity in hotspot regions may be detrimental to patient outcomes. Instead, we posit that transferring patients proactively out of developing hotspots to less affected regions, well before high ICU workload situations emerge, will improve overall patient outcomes.

## 1 Introduction

The coronavirus pandemic that started in late 2019 through the transfer of the SARS-CoV-2 virus to humans has been widely covered, and does not need further introduction. The ongoing fight against the recurring waves of this pandemic is a complex undertaking that has several key dimensions: prevention measures that include social distancing and personal protective equipment, such as masks; identification efforts via testing, tracing and isolating of those in contact with infected patients; improving treatment in terms of drugs and treatment procedures; protection via vaccination. The management of the healthcare operations providing the actual care to COVID-19 patients is often an afterthought, which in our view is a critical omission. As has been widely stated under the slogan ‘flatten the curve’, avoiding hospital overload situations is vital for the continued provision of high-quality care. Yet, where measures to prevent the inflow of new COVID-19 patients that would overwhelm the healthcare system have been exhausted, effectively managing the available healthcare system capacity becomes paramount. This is the focus of our investigation, and we posit that the management of available healthcare system capacity is a further critical aspect in the fight against the coronavirus pandemic.

In this paper, we study the management of intensive care unit (ICU) capacity during the first COVID-19 wave in the spring of 2020 in the most-affected regions in three European countries: Italy, the Netherlands and Germany. We observe great differences in patient outcomes, in line with a growing evidence base that ICU capacity is a major factor in COVID-19 mortality in hotspot areas (see Catena and Holweg 2020, Rocks and Idriss 2020, Bravata et al. 2021, Janke et al. 2021 and others). We thus further explore the role of regional ICU capacity shortages on patient mortality, and study the related capacity management aspects, such as efforts to increase regional ICU capacity, stoppage of standard operations, and relocation of patients out of hotspots. Our quantitative analysis of the first wave shows that a high ICU workload is linked to mortality for COVID-19 in a non-linear way. Our subsequent qualitative investigation elucidates the main challenges that hospitals face in effectively managing ICU capacity for COVID-19 care. Overall, our findings demonstrate the clear need to transfer patients early and proactively out of developing hotspots to less affected regions, if disproportional mortality is to be prevented. We conclude with a set of recommendations how hospitals, regions, and even countries can prevent such mortality by sharing timely capacity information and by virtually pooling available ICU capacity, in order to pre-empt regional ICU overload situations.

## 2 Context

Upon further considering ways to effectively manage available healthcare system capacity in the specific setting of COVID-19 care, two related, but distinct concepts emerge in the operations management literature: *capacity* and *workload*. The capacity of a healthcare system, by and large, denotes the system’s ability to treat patients. Load, on the other hand, refers to the number of patients treated as a fraction of the capacity at a specific point in time. Previous research often considered a single critical resource, such as number of beds, when measuring capacity (e.g., KC and Terwiesch 2009), or express it in terms of the maximum number of patients that can be treated (e.g., Kuntz, Mennicken and Scholtes 2015). Although the capacity of a healthcare system, such as the ICU, is not a constant, it usually does not vary greatly over time. It can be expanded to a certain extent when needed, for example during the yearly flu season, or briefly reduced, for example to provide staff with time to recover from a busy period.

The sources and management of capacity and workload in healthcare is being addressed in the operations management literature (e.g., Catena, Dopson and Holweg 2020, KC et al. 2020, KC and Terwiesch 2012). Studies here generally highlight the need for an effective management of the flow of patients through the healthcare system, and there is a broad consensus in academia on the negative impact of workload on the quality of care. For example, a patient is more likely to be discharged early in situations with a high occupancy of bed capacity in an ICU, which in turn increases the likelihood of readmission to the ICU at a later time (KC and Terwiesch 2012). In the context of cardiothoracic surgery, KC and Terwiesch (2009) show that sustained periods of increased workload can even increase the likelihood of mortality. These findings are congruent with recent COVID-19 specific studies at patient, region and country level that link ICU capacity and workload to patient outcomes.

At patient level, Bravata et al. (2021) study 8,516 COVID-19 patients in the US and link their clinical outcomes to the ICU workload during their stay. Although not consistent over time, they find that strains on critical care capacity was associated with higher mortality. Specifically, they observe significant increases in mortality developing beyond a 50% COVID-19 patient load at the ICU. At regional level, Catena and Holweg (2020) observed drastic differences in COVID-19 mortality in two neighbouring regions in Italy, identifying very high ICU workload levels as a key difference. Similar findings were made by Janke et al. (2021), who use American Hospital Association data to show that regions in the US with fewer ICU beds were statically associated with greater COVID-19 mortality. At national level, in comparison, the link between ICU capacity and COVID-19 mortality is not as clearly visible. Rocks and Idriss (2020) compare the outcomes of the pandemic in 33 countries and found some links between hospital capacity and mortality, albeit not statistically strong relationships. Similarly, Sen-Crowe et al. (2021) found no significant association between the number of ICU beds/100,000 and COVID-19 deaths in comparing 183 countries, and imply that other factors, such as the availability of supplies and personnel, may have a greater influence on the mortality of COVID-19 patients.

In this paper, we build on these studies by applying considerations on capacity and load to the response of healthcare systems to the first wave of COVID-19 infections. Specifically, we study the COVID-19 patient load on the ICU with a particular focus on the impact of high workload situations and how resulting adverse patient outcomes can be avoided. While ICU capacity and workload differ across countries, high workload situations have been the norm, rather than the exception – also before the pandemic. The coronavirus pandemic resulted in an unexpected and strongly varying inflow of a new group of patients in ICUs around the world. As a response, hospitals stretched their ICU capacity significantly, for example by adding extra beds or ventilators. Hospitals can typically also repurpose some of their readily available ICU capacity to manage increases in COVID-19 patients, for example by postponing less urgent surgical procedures. Most care for COVID-19 patients, however, comes on top of the regular inflow of intensive care patients. Even relatively low COVID-19 patient loads can thus lead to high workload situations in the ICU.

## 3 Method

### 3.1 Approach

The challenges posed by the coronavirus pandemic are complex and multi-faceted, thus easily constituting a ‘messy’ problem (Ackoff 1981). The management of the pandemic at regional, national and international level raises many fundamental debates. For one, it is not clear how to define ‘success’ in terms of the desired outcomes. While the preservation of life is an obvious metric, arguments have been made that measures undertaken to prevent the undue loss of life – for example through lockdowns – have led to the postponement of regular healthcare, and may thus have increased mortality for non-COVID-19 patients. Equally, the adverse effects on mental health and economic welfare have been frequently pointed out. These debates extend far beyond the scope of this study, and in fact, are likely to remain a point of debate for public health scholars for years to come.

For the purpose of this study, we adopt the number of COVID-19 deaths as the main metric of consideration and seek to understand the extent to which managing the ICU workload can contribute to improving the patient outcome in terms of survival. We acknowledge that the relationship between the COVID-19 deaths and ICU workload will not be quite that straightforward. The complexity inherent in managing healthcare system capacity during COVID-19 patient surges also means that a purely quantitative analysis is clearly too limited to support meaningful conclusions. Thus, we adopt a research design where quantitative analyses are complemented with perspectives from key stakeholders in the hospitals, as well as regional and national coordination centres.

In our analyses we have chosen to compare the worst affected regions (in absolute number of cases) in three European countries: Lombardy (Northern Italy), Noord-Brabant (Southern Netherlands) and Bavaria (Southern Germany). While there are idiosyncrasies embedded in each national context, studying three different countries does provide the ability to extrapolate general patterns across the three contexts.

### 3.2 Data

We focus our quantitative analyses on the first coronavirus wave in Europe, which began around the end of 2019, but saw its main impact during March and April 2020. The coronavirus pandemic is arguably one of the most documented healthcare events of all times, with many institutions tracking case prevalence, recuperation and mortality rates in near real-time. Most prominently these include John Hopkins’s COVID-19 dashboard, the COVID-19 tracking project in the US, and of course, the dashboards of the WHO, the EU, as well as a series of national agencies.

For the purpose of this study, we retrieved data on COVID-19 deaths from the Italian Civil Protection Department, the National Institute for Public Health and the Environment in the Netherlands (RIVM) and the Robert Koch-Institut (RKI) in Germany. We acknowledge that this metric is far from perfect. Even though patients who are dying are more likely to be tested than asymptomatic patients, the reported number of COVID-19 deaths can be distorted by differences in the level of testing, time lags in reporting and divergence in the respective national definition of what constitutes a COVID-19-related death. To control for these limitations, we compared the reported COVID-19 deaths with data on ‘excess deaths,’ that is, deaths occurring above the long-term stable level. During the first wave, excess deaths will primarily include COVID-19 deaths, but it must be acknowledged that excess deaths may also stem from the potential postponement of other critical care due to the focus on COVID-19. Equally, the improved hygiene measures and reduced accident deaths due to social distancing and national lockdowns are likely to reduce non-COVID mortality. Despite these potential limitations, we found that in the regions we studied COVID-19 deaths was highly correlated with excess deaths^3^, which would suggest that the impact of these potentially distorting factors is limited.

We study the relationship between COVID-19 deaths and ICU workload, specifically, the load of COVID-19 patients as a fraction of the total number of ICU beds available at that point in time. To this end, we employed data about the number of COVID-19 patients occupying an ICU bed and the total number of ICU beds available in a region per day. For Italy, we use data from the Italian Civil Protection Department and Lombardy’s regional government; for Germany, we received data from the German Association of Intensive and Emergency Care (DIVI); and in the Netherlands we received data from the national coordination centre for transferring patients (LCPS). It must be noted that these data are self-reported by hospitals in each region and thus may be subject to interpretation of the individual providing the data. Regional comparability is hindered by the lack of a straightforward definition of an ‘available’ ICU bed. In general, it will simply refer to the number of ICU beds that can provide intensive care (with monitoring and support functions) for a COVID-19 patient 24 hours a day, every day of the week.

### 3.3 Interviews

Given the limitations inherent in any COVID-19 related data, it became quickly apparent that further qualitative data was needed. Hence, we conducted interviews with key stakeholders in each of the regions to help us understand and put into context the quantitative analysis. In total, we conducted 21 interviews for the purpose of our study. As shown in Table 1 below, the interviews were done primarily with critical care physicians and hospital managers, but also with the staff and managers of coordination centres for transferring patients that were put in place in Germany and the Netherlands. Despite being in the midst of the largest (public) health crisis in recent history, these stakeholders were willing to share their experiences and views on the matter. The length and format of the interviews varied greatly, however, depending on the time the interviewees were available to us.

**Table 1:**
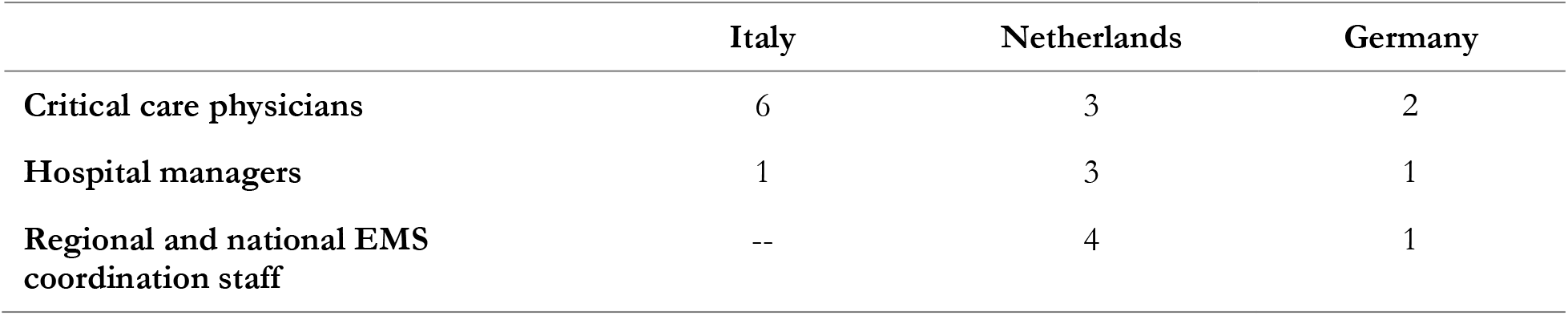
Overview of the interviews conducted, by function and country.

## 4 Case findings

In this section, we briefly discuss the course the pandemic took in each of the three regions in our study, before bringing case findings together in a cross-case analysis.

### 4.1 Northern Italy: Lombardy

Lombardy is one of the twenty administrative regions within Italy and was the Italian hotspot of the COVID-19 infections in the first wave. Lombardy is a wealthy region that is home to 10 million inhabitants and the centre of industrial production in Italy. According to OECD data^4^, prior to the COVID-19 outbreak, Italy had an ICU capacity of 8.6 per 100,000 inhabitants.

Lombardy identified the first positive COVID-19 case on February 20, 2020. The patient was a 38-year-old man with a pneumonia that was not responding to treatment. Within 24 hours from the first positive case, 36 other cases were identified, and concerns started to rise about a cluster with an unknown number of positive COVID-19 patients (Grasselli, Pesenti and Cecconi 2020). The first death was on February 21 and, in one week, the total number of positive cases had risen to 615 and there were already 80 ICU COVID-19 patients.

The first wave of COVID-19 in Lombardy was a tsunami of patients no-one in the region was expecting, with a disease that was still not well known. Lombardy was the first region in Europe to experience the COVID-19 crisis. The knowledge health care professionals had about the virus was limited to the information coming from the initial hotspot in Wuhan. Hospitals were expecting to treat patients with a respiratory disease, but instead COVID-19 manifested itself as a disease with numerous possible complications including kidney failure, thrombotic complications, stroke and myocardial infarction. Clinicians had to quickly learn how to treat the new disease while more and more patients were arriving in the emergency departments. In the 548-beds hospital Ospedale di Cremona, by the end of February, around 100 COVID-19 patients were arriving in the emergency department every day.

Lombardy acted quickly to increase the ICU capacity. From the onset of the epidemic, the number of ICU beds in Lombardy increased from 7.2 to 15.9 beds per 100,000 inhabitants. The response of the hospitals was coordinated by the regional government with the support of the national government through the Protezione Civile, the Italian body managing national emergencies. There were two weekly webinar meetings organised by the Centro di Coordinamento Regionale delle Terapie Intensive, the regional team in charge of coordinating the ICU resources in the regional network. The hospitals were having daily calls with the team to update them on the availability of resources and on what they needed. There was a formal policy of hospital transfers within the region, but between March 7 and April 4 only 78 patients were actually transferred to other regions (Paolini 2020).

Despite the containment measures, the increase in the number of ICU patients was relentless. The peak of new positive cases was reached on March 21 and the first time the number of ICU patients started to decrease was only on March 31 when there were 1,324 patients in the ICUs in the region. By June 30, 2020, the Italian Civil Protection Department reported that Lombardy had experienced 165 deaths per 100,000 inhabitants.

### 4.2 Southern Netherlands: Noord-Brabant

Noord-Brabant is one of 12 provinces within the Netherlands. With 2.5 million inhabitants, it is the third-largest province of the Netherlands in terms of its population and economy. According to OECD data^5^, prior to the COVID-19 outbreak the Netherlands had an ICU capacity of 6.7 beds per 100,000 inhabitants.

The first case was identified on February 27, 2020 in a hospital in Tilburg, and the first death was reported on March 8, 2020. Around the same time, hospitals in the region started sounding the alarm; they postponed most surgery and tightened visiting regulations. By the end of March about 70% of all ICU beds in the region were in use for COVID-19 care. From then on, several hospitals faced serious capacity issues. The region has 10 hospitals totalling 4.7 ICU beds per 100,000 inhabitants prior to COVID-19, which at the peak of the first wave was increased to 10.9 ICU beds per 100,000 inhabitants. Noord-Brabant was the Netherlands’ hotspot of the COVID-19 infections in the first wave. Between Feb 27 and June 30, 2020, the region saw 2,777 hospitalizations with COVID-19 (108.3/100,000 inhabitants), and 60.0 deaths per 100,000 inhabitants with COVID-19 being the known cause.

The overflow of ICUs in the region led to the transfer of many patients to other – less affected – regions in the Netherlands and Germany. Between early-March and the end of June a total of 900 patients were transferred out of the region, over 600 of which during the surge of COVID-19 patients in March. First, this was organized bilaterally between ICU doctors of individual hospitals. Quickly, the national government initiated the instalment of a national coordination centre overseeing and managing the transfer of patients. This coordination centre was operational by March 27. The national coordination centre communicates with 12 regional coordination centres who coordinate among the hospitals in their region. Hospitals in Noord-Brabant report their capacity and patient numbers to the regional coordination centre. The regional coordination centre supports the transfer of patients from one hospital to another in the region or requests transfers out of their region to other regions via the national coordination centre if needed.

Preparing for the worst, the national and regional governments also focused on increasing the health system’s capacity. At a national level, the goal was to quickly ramp up from 1,150 to 2,400 ICU beds. This was mostly achieved by postponing regular care and bringing all available resources to the ICU. The national government orchestrated the purchasing of 4,000 ventilators from several suppliers across the world. Several regional governments initiated several emergency hospitals and care facilities, with convention centres MECC, in Maastricht (capacity: 276 hospital beds) and Ahoy, Rotterdam (initial capacity: 82 beds and a maximum of 680) being the largest facilities.

### 4.3 Southern Germany: Bavaria

Bavaria is one of Germany’s sixteen federal states, and with a population of 13.8 million it is the most populous one. The region also has the highest number of healthcare facilities in Germany, 548 in total. The ICU capacity of the region was 28.7 per 100,000 inhabitants prior to the COVID-19 pandemic, which was ramped up to 35.3 at the peak of the first wave (LDL, 2020). In comparison, pre-pandemic Germany overall had a considerably higher ICU capacity (33.9 beds per 100,000 inhabitants), compared to Italy (8.6) and the Netherlands (6.7).

Bavaria was also the starting point of the COVID-19 pandemic in Germany, which infected 48,382 and led to 2,591 deaths by June 31, 2020. The first German COVID-19 case was discovered January 27 2020 in Starnberg. The vector of infection was an attendee at an internal seminar at car parts supplier Webasto. The attendee infected 13 co-workers prior to falling ill on the return flight to China. It took another month until the next set of cases were discovered, from which point onwards the pandemic started developing. In some ways this one month was helpful in preparing the response.

As Germany is a federal republic, the responses varied considerably by federal state. This included timing and severity of lock-down measures, as well as regional policy measures. Also, emergency services are under state control, including the police. The federal state initiated the development of a national system to record ICU bed capacity. As part of the national registry of ICU capacity (named the ‘<u>DIVI registry</u>’) as it is administered by the German association for intensive care and emergency medicine (DIVI), all hospitals in Germany were required to report on their available ICU capacity every morning. The DIVI registry was set up by March 27 and it became mandatory for all hospitals to submit a daily report by April 9. As part of this report, both actual capacities as well the so-called ‘7-day emergency capacity’ had to be reported. The latter refers to the additional ICU bed capacity hospitals could install with a 7-day notice period. There have only been isolated instances of transferring patients outside their regions during the first wave, although this changed drastically during the second wave in late 2020/early 2021 as several regions reached critical capacity utilisation levels, when pan-regional transfers became widely used.

#### 4.4 Cross-case analysis

To provide an overview of the respective ‘waves’, Figure 1 plots the course of known COVID-19 cases per day across the three regions. As can be seen, there is a clear pattern that replicates across all regions. We see in Figure 1 that Lombardy was the first region to see exponential spread of the coronavirus. The outbreaks in Noord-Brabant and Bavaria largely coincided but lagged the outbreak in Lombardy both in timing and scale.

**Figure 1:**
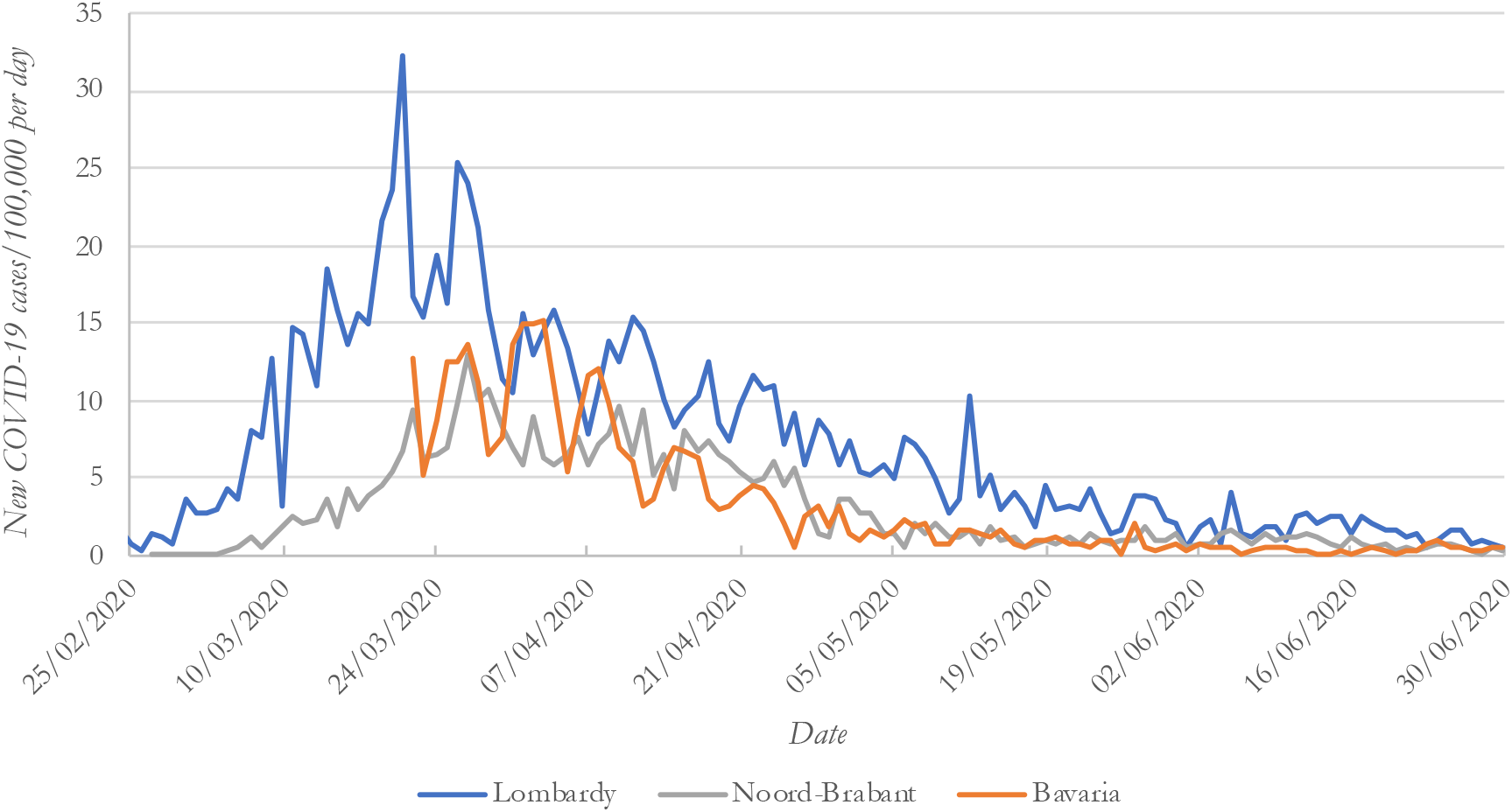
Newly reported COVID-19 cases per day, by region.

COVID-19 deaths match the pattern of newly reported cases, albeit with a delay, as shown in Figure 2. As the timing of the testing – and thus confirmation of infection – is not conducted at a standardised point in time, there is no direct correlation between the reported cases and mortality. Indeed, we theorize that the rate of mortality is partly a function of the healthcare system’s capacity to treat patients to the same standards, which in turn determines the different patient outcomes for Lombardy, Noord-Brabant and Bavaria. In the following we try to elucidate the complex mechanisms how ICU capacity and workload link to COVID-19 mortality.

**Figure 2:**
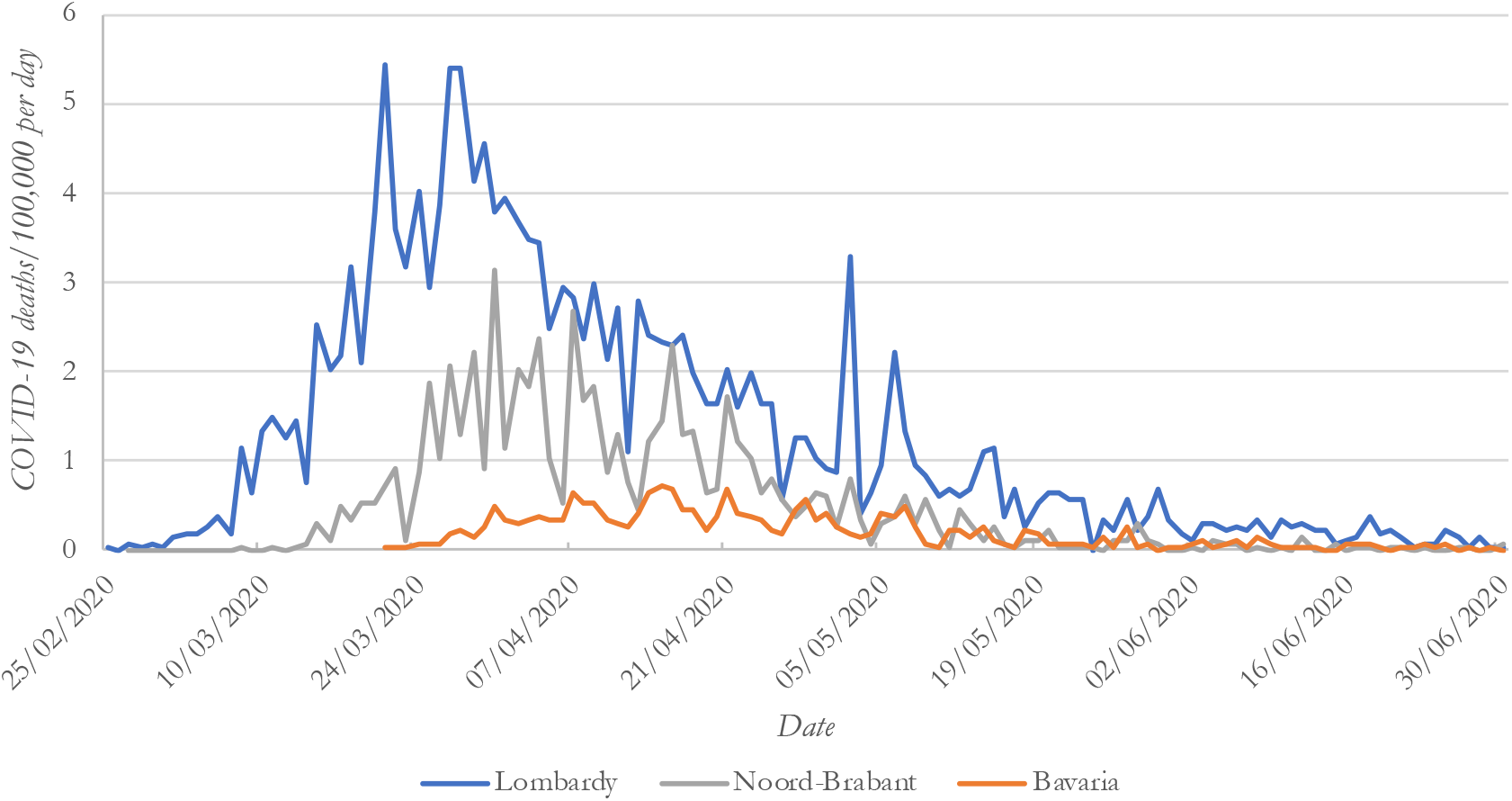
COVID-19 deaths per day, by region.

### 4.5 The effects of ICU overload on patient mortality

By merging the data from the three regions, we can assemble an empirical picture of how COVID-19 patient load on the ICU and COVID-19 deaths link, as shown in Figure 3. Here, one would expect that mortality rises linearly with the COVID-19 patient load as a fraction of total ICU capacity at that point in time. Yet, COVID-19 deaths clearly rise non-linearly. In fact, one can approximate this non-linear relationship to a high degree (R^2^ = 0.88) with a quadratic equation (y = 6.53x^2^ – 1.19x + 0.08).

**Figure 3:**
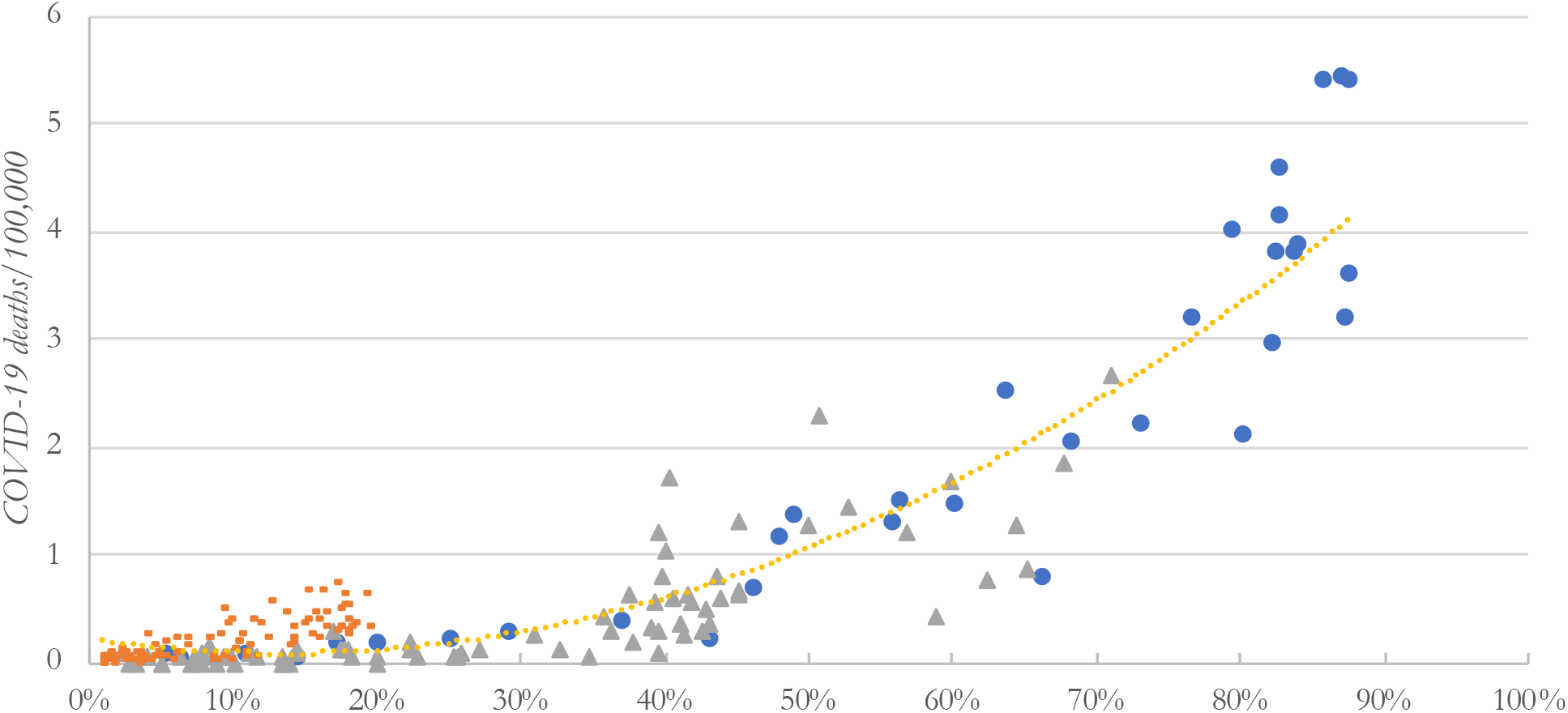
COVID-19 deaths as a function of COVID-19 patient load on the ICU.

It is notable that the sharp increase in COVID-19 deaths already starts at relatively low COVID-19 patient load on the ICU, which can be explained by the fact that there are also other patients admitted that require intensive care. The exponential rise in mortality is clearly driving the high mortality in Lombardy, compared to the very low mortality in Bavaria. Noord-Brabant, in comparison, sees both the low mortality like Bavaria in situations of low ICU loads, yet replicates the high mortality experienced in Lombardy when confronted with higher ICU loads. Our finding confirms the observations reported by Bravat et al. (2021), who also found that strains on ICU capacity lead to an increase in mortality once the load of COVID-19 patients on the ICU exceeds 50% of total ICU capacity, and to a very high increase beyond a 75% load.

The trendline shown in Figure 3 has an unmistakable resemblance to Kingman’s formula (1961) that discusses how queues form in a single-server line. This becomes intrinsically logical when we clearly distinguish between capacity and workload. As the number of COVID-19 patients admitted to the ICU quickly increased, hospitals in the regions responded by expanding the number of ICU beds. In Lombardy and Noord-Brabant, the added capacity was quickly occupied by newly arriving COVID-19 patients and the COVID-19 patient load started to approach the total maximum ICU capacity^6^.

It is also possible to explain our empirical findings analytically by using a correction factor to represent the decreasing marginal benefits of increasing the number of beds. In the context of care for COVID-19 patients, we define and measure workload as the fraction of COVID-19 patients and the total number of ICU beds available at that point in time. In this fraction, the numerator increases as more COVID-19 patients are admitted. The marginal benefits of additional ICU beds, however, decrease because other critical resources cannot keep up. This helps explain the non-linear increase in mortality as the COVID-19 patient load on the ICU ward increases. To this effect we can define a correction factor *c*(*t, u*) as a monotone and strictly decreasing function of the number of COVID-19 ICU patients *t*, and the number of ICU beds *u*. We then can define mortality as a function *z*(*t, u*) = ß · *w*(*t, u*) + *const*., where ß is linear coefficient of the daily mortality and *const*. a constant. By adding the correction factor *c*(*t, u*) into this function, we get:

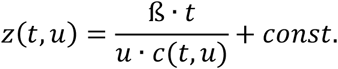

Assuming a specific monotone strictly decreasing function, such as for example

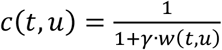

with constant *γ*, and where we assume that *w*(*t, u*) is monotone strictly increasing, this provides us with the following function for the mortality:

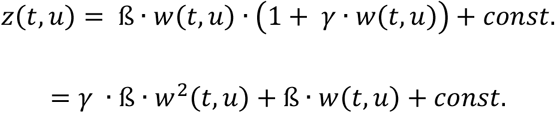

Thus, assuming that the marginal benefits of additional ICU beds decrease, it is conceptually straightforward to show the resulting non-linear rise in patient mortality as the COVID-19 load increases on the ICU.

## 5 Discussion

We set out to investigate how the management of ICU capacity can improve outcomes for COVID-19 patients. Three important implications emerge from our analyses: first, the notion of ‘capacity’ is being interpreted too simplistically in this context; second, we show that a rising COVID-19 patient load leads to a disproportionate deterioration in patient outcomes; and third, we will argue that proactive capacity management can help avert these adverse outcomes. We will discuss each aspect in turn.

### 5.1 A systems view of ICU capacity

The first, and arguably most profound, implication from our study is that the way how the concept of ICU ‘capacity’ is being used in the present discourse is not only misleading, but in fact substantively contributing to adverse patient outcomes. Looking back, when the demand on the healthcare system rose during the first wave, the attention universally turned to the notion of the number of ICU beds available to deal with COVID-19 patients in the first instance. Later on, the number of available ventilators became the focus of public attention. During the second wave, staff shortages due to sickness and exhaustion became prevalent. In Germany, for example, Uwe Janssens, president of the German Association for Intensive Care and Emergency Medicine (DIVI) estimated, there is a nationwide shortage of between 3,500-4,000 ICU nurses in Germany. He stated that ‘we have a dramatic shortage of nursing staff. By now we have sufficient capacity in terms of ICU beds and ventilators. But those alone do not help if we don’t have the staff to look after the patients’ (ARD 2020). What this change in discourse highlights is that the notion of capacity is much more complex than simply counting any physical artifacts.

Previous operations management research often considered a single critical resource in a ward, such as number of beds, when measuring capacity (e.g., KC and Terwiesch 2009). Although this type of measurement is typically a good proxy of the supply of care a ward can provide, in the context of COVID-19, multiple factors, such as availability of space on the ward to provide treatment for a patient (the ‘bed’), physical equipment like mechanical ventilators, and trained medical staff, all need to be taken into consideration to assess the overall capacity. Increasing the number of ICU beds and ventilators, without increasing the other resources required to provide care does not necessarily result in a proportional increase of capacity. Other research considered the maximum number of patients treated as a measure of capacity (e.g., Kuntz, Mennicken and Scholtes 2015). This measurement of capacity has the benefit to consider how much resources can be stretched to serve the inflow of patients; however, it is an endogenous variable. In the context of the coronavirus pandemic, an increase of capacity defined in these terms does not necessarily imply that the extra patients are treated at the same level of care as if the number of patients was lower.

Changing the conceptualisation of ICU capacity from a single resource, like beds or ventilators, to a systemic view that encompasses the physical artefacts (beds, ventilators and other equipment), labour (in terms of doctors, nurses and other support functions), and the embeddedness with the wider system (ambulances, telemetry, laboratory and other examination services) is the essential first step. We need to understand intensive care as a system comprising many subsystems. The ICU itself is embedded within a wider healthcare system: to provide critical care, the physical artifacts like ICU beds, the medical equipment like ventilators, and human resources are needed. This infrastructure needs to be embedded within the hospital system, for example, to provide technical gases (e.g., oxygen), as well as telemetry and examination services, like laboratory and radiology services. The hospital system is embedded in a wider healthcare or even welfare system including, for instance, capacity in nursing homes for final recovery, ambulances to transfer patients and palliative care.

Taking a system’s view of intensive care provision leads to an intuitive yet important conclusion, namely that the level of care provided is ultimately a function of each of the subsystem’s availability. This has important ramifications for the increase of ICU capacity. The faster capacity is increased, the greater the likelihood that not all resources can be increased at an equal rate. ‘An essential aspect was the staff. Increasing the number of ICU doctors when you increase ICU beds is a significant challenge’ (interview with a physician from Italy). The implication on the quality of care was clearly visible during the COVID-19 pandemic, as experiences in the Netherlands. ‘It is important to work with a kind of lower quality limit [to avoid] treating patients in such a manner that they do not really stand a chance […]. What you see is that in healthcare everyone keeps on going, even below that limit. That is what nurses are saying today: the quality of care has been at stake. Looking back, that’s only logical’ (interview with a physician from the Netherlands). Put differently, as constraints on any aspect of the care system, be it ventilators or trained ICU nurses, emerge, the quality of care will suffer.

### 5.2 The impact of increased COVID-19 patient load on the ICU

An important empirical finding of our study is that mortality increases exponentially as the COVID-19 patient load on the ICU increases – as distinct from a linear, or near-linear increase that one could expect. Building on existing research that has uncovered similar relationships between workload and patient outcome (e.g., KC and Tierwisch 2009), we propose the following explanation for this phenomenon.

A common response to regional surges in the first wave of the coronavirus pandemic was to increase regional ICU capacity. This was done mostly by reassigning medical staff from other departments to the ICU and shutting down routine operations in order to free up ICU capacity for COVID-19 care, but also by opening entirely new wards and even hospitals (e.g., the Nightingale hospitals in the UK). The training of ICU medical staff, however, cannot be accomplished in such a limited amount of time and the staffing of these structures can become a major issue. ‘It takes years of training to become a specialist. When you are a specialist, you are a junior specialist, to become a skilled specialist you will need more years’ (interview with a physician from Italy). Once COVID-19 patients need intensive care, they are very labour-intensive to care for. The patients themselves require many treatments and their situation can deteriorate very quickly (the so-called ‘cytokine storm’), which means patients need to be monitored very closely. All medical staff that is working with COVID-19 patients has to do so in full PPE, which makes work exhausting as heat builds up under the gown and mask.

Our empirical data shows that in regions where the number of ICU beds was drastically increased, hospitals struggled with increasing other resources, most notably ICU nursing staff. In Lombardy, for example, the healthcare system was stretched by increasing the number of beds, yet to a point where health care professionals with a different specialization had to be retrained to support COVID-19 care. ‘The recruitment efforts were directed to the anaesthesiologists. The most experienced helped the ones who had not been in reanimation for some time. This was also true for nurses. Every nurse with some competence in the ICU was recruited, like operating room nurses. They were assigned to new tasks and to a new workplace. These people were incredibly willing to help and had a lot of passion and dedication. They worked for 14-16 hours per day without complaints’ (interview with a physician from Italy).

New recruitment efforts were moderated by rising staff absenteeism levels, either from falling ill by COVID-19 themselves, or from the enormous physical and psychological strain placed on them by caring for their patients. Staff absenteeism only puts further pressures on the remaining medical staff. At the height of the first wave in the UK, critical care nurses were asked to look after four to five patients, compared with one or two during normal operations. Many intensive care physicians have warned of the resulting effects on the quality of care, such as Simon Walsh, an emergency medicine consultant and British Medical Association representative, said the secondments would inevitably stretch patient-staff ratios elsewhere. ‘We understand the need (..) when we’re deploying doctors that are already providing care elsewhere, the ratio of doctors caring for critical care patients is going to reduce’ (Financial Times 2020).

### 5.3 Avoidance of regional surges by proactively transferring patients

While attempts to increase regional ICU capacity in response to regional surges in COVID-19 patients was a logical response, our findings suggest this approach is based on the wrong way of thinking about capacity. Instead, we argue that transferring patients out of regional hotspots is key to reducing the mortality rate. Transfers must start early and proactively, and long before the COVID-19 load exhausts regional ICU capacity. This finding goes counter to common healthcare practice and the ethos of critical care physicians to treat patients, not sending them away. Yet, our empirical findings highlight what critical role the transfer of COVID-19 patients can play. These findings are in line with the longstanding appreciation in the operations management literature for how networks can reduce the risk of supply disruption by pooling capacity (e.g., Kim, Chen and Linderman 2015). In the context of COVID-19 there are early examples too of how network-based strategies have been employed successfully by some regions to manage patient admissions (Pett et al. 2020).

All regions attempted to facilitate a proactive transfer of patients, albeit with very different organizational structures and with varying success. A key impediment for transferring patients is the need for transparency about ICU capacity across the country. Staff of hospitals and the coordination centre in Noord-Brabant explained that, despite the general willingness of other regions to take over patients from, they were struggling to actually find operational ICU beds in less-affected regions in the Netherlands, at some point transferring patients to Germany. This struggle with identifying available ICU beds is still hindering the coordination centres in the Netherlands during the third wave to this day.

Germany provides a good example on how to make ICU capacity transparent, and use that transparency to avoid regional overload situations. As the first wave unfolded, the German association for intensive care medicine (DIVI) jointly with the Robert Koch-Institute (RKI, Germany’s national public health administration) established a web-based registry of ICU bed availability. Patient transfers within Germany were the exception during the first wave yet became increasingly more common during the second wave. As Saxonia developed into a hotspot area, for example, patients were increasingly flown out of the Bautzen and Görlitz counties to major hospitals in Dresden and Leipzig, but also as far as Brandenburg. These transfers are coordinated by the regional emergency service coordination centres. The DIVI registry does demonstrate that digital technologies, coupled with political will, can provide near real-time capacity information in support of a proactive capacity management system that prevents regional overload situations. The DIVI registry has not been without its criticisms, largely related to its failure to integrate fully with existing hospital bed management systems in operation in several federal states, yet in our view denotes a best practice case how to address a critical public health crisis. Even in the absence of a formal IT infrastructure, any regional coordination mechanisms can significantly reduce any excess mortality of COVID-19 patients where the workload of ICUs increases.

Case evidence that patient transfers can be of great advantage is already being reported: during the second wave, London was emerging as a particular hotspot in the UK, adding great strain to all London hospitals. Initially, ICU bed capacity in London nearly doubled, yet this was not found to be sufficient. The ‘pandemic transfer service’ was then instigated in order to distribute patients to regions with lower COVID-19 caseloads. In total, 2,300 patients have been moved between September 2020 and March 2021, in some cases up to 300 miles outside of London. Overall, about one in eight COVID-19 patients admitted to an ICU in London during the second wave had to be transferred to another region (BBC 2021). Similar measures have been taken by other countries too. Italy, for example, has introduced a train-based patient transfer service that provides the ability to transfer COVID-19 patients efficiently across regions.

## 6 Outlook

The need to manage the capacity of healthcare systems has been highlighted in the operations management literature, and the challenges emerging with the COVID-19 pandemic mark a case in point. The workload of regional ICUs in particular has been identified as a critical factor in explaining patient mortality (Catena and Holweg 2020, Rock and Idriss 2020, Bravata et al. 2021, Janke et al. 2021). Our study adds a critical empirical aspect to this debate, demonstrating that mortality is likely to increase non-linearly as the COVID-19 patient load rises on the ICUs.

Our findings and recommendations have immediate managerial relevance as many countries face the challenges of more virulent strains of the SARS-CoV-2 virus, as well as the long-term prospect that further mutations will turn COVID-19 into a recurrent seasonal disease (Murray and Piot 2021). We do know from the experience to date that commonly regions are not affected equally, but that regional hotspots tend to emerge at different times within countries. Our main recommendation thus is to stop stretching local ICU capacity. Instead, patients should be transferred early and proactively out of emerging hotspots to less affected regions, in order to prevent the non-linear rise in patient mortality as the COVID-19 workload on regional ICUs rises. We base this recommendation on our empirical evidence, yet our conclusion is supported both by early modelling evidence (Alban et al. 2020, Lacasa et al. 2020), as well as case evidence from regions that have successfully employed a patient transfer strategy (Pett et al. 2020).

It is clear that much remains to be learnt about how to deal with COVID-19, yet as our findings show, the pan-regional management of our healthcare systems’ capacity is another important tool in our fight against this pandemic.

## Data Availability

All data used for the quantative part of the paper is available in the public domain. References are provided in the paper.

https://coronavirus.jhu.edu/map.html

## Footnotes

3 The positive correlation between excess deaths and mortality was 0.92 for Lombardy (Italy), 0.96 for Noord-Brabant (the Netherlands) and 0.89 for Bavaria (Germany).

4 Beyond Containment: Health systems responses to COVID-19 in the OECD (April 16, 2020).

5 Beyond Containment: Health systems responses to COVID-19 in the OECD (April 16, 2020).

6 Specifically, we find a positive correlation between ICU capacity and workload of 0.92 in Lombardy and of 0.86 in Noord-Brabant. As Bavaria did not increase capacity, measuring the correlation between ICU capacity and workload is not relevant.

## Notes

### Competing Interest Statement

The authors have declared no competing interest.

### Author Declarations

Approval for conducting the interviews has been granted by the University of Groningen' Institutional Review Board under reference #FEB-20200813-11644.

